# MuSTAF: Clinically Relevant Multi-task Spatiotemporal Attention Fusion Framework for Breast Cancer Detection with Longitudinal Mammography

**DOI:** 10.64898/2026.07.07.26357474

**Authors:** Yutong Li, Austin Castelo, Jennifer B. Dennison, Nicole M. Kettner, Weiva Sieh, Joshua R. Joseph, Edward Castillo, Kristy Brock, Olena O. Weaver, Chengyue Wu

**Author notes:** This work has been submitted to the IEEE for possible publication. Copyright may be transferred without notice, after which this version may no longer be accessible.

## Abstract

Recent NCCN guideline highlighted AI-based mammographic risk prediction, but AI-based breast cancer detection remains questionable to translation. One barrier is current models often do not match routine clinical reasoning, which may add decision burden than benefits. In practice, radiologists compare current and prior mammograms while assessing breast density, bilateral symmetry, and lesion laterality. To align AI with this reasoning, we developed MuSTAF, a multi-task spatiotemporal attention fusion model for patient-level breast cancer classification from longitudinal full-field digital mammography. MuSTAF uses up to three recent mammograms, integrates temporal and cross-view information, refines suspicious-region features, and jointly predicts cancer status, breast density, and bilateral symmetry, with a separate laterality classifier for cancer-positive cases. In an internal case-control cohort (n = 351), MuSTAF achieved a cancer classification (AUC=0.84) exceeding all architecture-level baselines and published mammography AI models adapted to the same task (AUC ≤ 0.81). Simultaneously, it achieved AUCs of 0.83/0.80 for density/laterality assessments, and removing these auxiliary tasks reduced cancer detection performance. On the external CSAW-CC dataset (n = 8,723), model performance improved from 0.72 to 0.88 when restricting cancer cases to those with latest exams within 60 days before diagnosis, showing that temporally distant labels may shift detection evaluation toward risk prediction. Longitudinal analysis further showed that three recent exams outperformed five exams internally (AUC = 0.84 vs 0.80) and externally (0.72 vs 0.66), indicating recent imaging evidence mattered more than remote history. Overall, MuSTAF model improved longitudinal mammographic cancer classification while providing auxiliary outputs, and clarified temporal factors for applying AI to screening detection.

## I. Introduction

BREAST cancer is one of the most common cancers worldwide. In the United States, it accounts for nearly 30% of cancers among women [1]. Early detection through screening and timely intervention has significantly reduce breast cancer mortality [2]. Among available screening modalities, mammography (MG) is one of the most widely used because it is accessible, cost-effective, and provides high spatial resolution with relatively low radiation dose. However, MG can miss cancers because of overlapping fibroglandular tissue, subtle lesion appearance, reader fatigue, and the complexity of bilateral multi-view interpretation [3]. Computer-aided detection and diagnosis (CAD) systems have been developed to assist radiologists in screening MG [4]. Recent AI studies have further improved screening performance, including reduced false-negative findings and reduced screen-reading workload [5, 6].

Despite this progress, many deep learning models for early cancer detection from screening still questionable regarding clinical translation. The major barrier is current models often do not match routine clinical reasoning, which may add decision burden than benefits to clinicians. In particular, first, radiologists evaluate breast density, bilateral symmetry, and lesion laterality in routine MG interpretation for cancer detection. Breast density is part of BI-RADS reporting and is associated with both masking and cancer risk. Bilateral asymmetry is an important cue for detecting unilateral or developing abnormalities. Tumor laterality provides an anatomical localization output for interpretation and quality control [7]. Conversely, many MG-based AI systems directly provide a “blackbox” cancer/non-cancer category or probability. These outputs, even if having a high overall performance, often lack clinically relevant intermediate information as uncertainty checkpoints, which undermines radiologist trust and limits error review [8]. These factors suggest that clinically relevant multi-task learning has great potential to improve both performance and interpretability [9-11]. Second, in clinical practice, radiologists often compare current and prior mammograms to identify new or changing findings. Longitudinal information is especially important in screening, where temporal changes can provide substantial cues for ruling out false positive lesions with low risk. However, most CAD/AI models analyze a single screening examination in isolation [12]. Recent studies have explored recurrent networks, temporal attention, and transformer-based encoders for longitudinal MG analysis [13-16]. However, few models integrate longitudinal modeling with clinically meaningful auxiliary outputs in a unified framework.

In this study, we developed a Multi-task Spatio-Temporal Attention Fusion model (MuSTAF) for individual-level breast cancer classification from longitudinal MG. MuSTAF integrates serial bilateral CC and MLO views, temporal fusion, suspicious-region refinement, breast density classification, bilateral symmetry learning, and tumor laterality assessment. We evaluated MuSTAF using patient-level stratified 7-fold cross-validation in an internal screening cohort and external validation on the public CSAW-CC dataset [17]. This study assesses whether clinically relevant longitudinal multi-task modeling can improve cancer classification while providing interpretable auxiliary outputs for mammographic screening.

## II. MATERIALS AND METHODS

### A. Study Design and Cohorts

The internal model development and validation leveraged a prospectively collected screening data from the MERIT (Mammography, Early Detection, Risk Assessment, and Imaging Technologies) cohort, acquired between 2017 and 2021, under approval of the MD Anderson Cancer Center Institutional Review Board (ClinicalTrials.gov identifier: NCT03408353) [18] with a waiver of informed consent. As shown in **Table 1**, the in-house cohort included 351 selected from the MERIT cohort using a nested case-control design (176 breast cancer cases and 175 cancer-free controls, including 67 benign and 108 negative cases). Each participant contributed an average of 3.8 serial annual examinations. Breast density from each examination was extracted from the corresponding radiological reports and dichotomized as non-dense (BI-RADS A–B) versus dense (BI-RADS C–D) [19].

**TABLE I.** Patient Distribution in Used Dataset.

| Category | Total | Breast Status |  | Breast Density |  |
| --- | --- | --- | --- | --- | --- |
|  |  | Cancer | Non-Cancer | Dense | Non-dense |
| In-house | 351 | 176 | 175 | 185 | 166 |
| CSAW-CC | 8723 | 873 | 7850 | 673 | 8050 |

External validation used the public CSAW-CC MG dataset from Karolinska University Hospital, Stockholm, Sweden [17]. The non-hidden subset included 873 women with first-time breast cancer and 7850 healthy controls, with an average of 2.8 examinations per individual. External breast density labels were derived from LIBRA quantitative density measures [20].

#### B. Image Processing and Model Training

Each examination included four standard full-field digital MG views: bilateral craniocaudal (CC) and mediolateral oblique (MLO) images. Images were converted from DICOM to PNG after modality and Volume of Interest (VOI) lookup table transformations. Pixel intensities were normalized using foreground percentile-based normalization and contrast-limited adaptive histogram equalization [21]. Images were then resized to 224 × 224 pixels. To construct examination-level inputs, right breast images were horizontally flipped, then the bilateral CC views and bilateral MLO views were each concatenated horizontally and combined vertically into a single composite. Longitudinal sequences were ordered chronologically, retaining the three most recent examinations up to and including the latest screening before diagnosis (cases) or the last available screening (controls) per individual. Sequences shorter than three were left-padded by repeating the earliest examination, with a padding mask provided to the temporal attention modules to distinguish padded from true time points.

Models were implemented in PyTorch [22] and trained on NVIDIA Tesla A100 GPUs. MuSTAF was optimized using a multi-task objective that combined cancer classification loss, breast density classification loss, and symmetry regularization. Learnable SoftMax-normalized task weights were used to adaptively balance the objectives. The independent laterality classifier was trained separately using cancer cases with laterality labels. Its output was used only as an anatomical localization and quality-control output during inference.

Internal validation used individual-level stratified 7-fold cross-validation. All examinations from the same individual were assigned to the same fold to prevent data leakage. The AdamW optimizer [23] was used with a learning rate of 1 × 10−^4^, weight decay of 1 × 10−^4^, and batch size of 4. Models were trained for up to 100 epochs with early stopping based on validation cancer AUC (Area Under the Curve). For external validation, the seven fold-specific MuSTAF models were applied to CSAW-CC. Predicted probabilities were averaged across folds to generate the final ensemble prediction.

### C. Model Architecture

We developed MuSTAF to jointly learn cancer-discriminative features, density-related representations, and bilateral asymmetry patterns from longitudinal multi-view MG (**Figure 1**). For a longitudinal sequence with ***T*** examinations, the model input is denoted as: 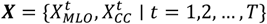, where 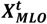 and 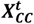 denote the bilateral CC and bilateral MLO composite images at time point ***t*** after left-right view alignment and concatenation.

**Fig 1.**
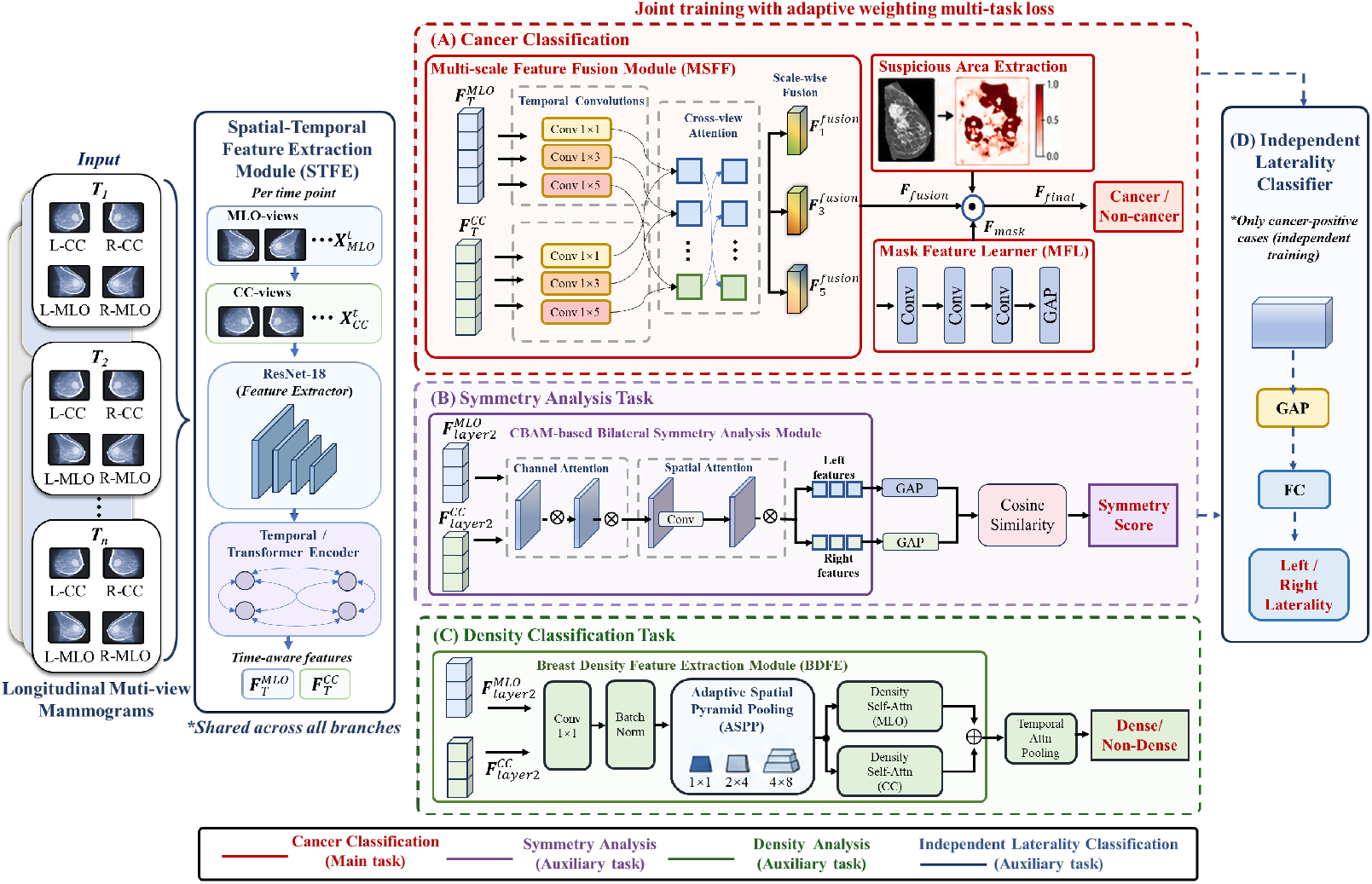
Overall architecture of the proposed MuSTAF framework for multi-task longitudinal MG analysis. Longitudinal mammograms acquired at multiple screening time points with four standard views (L/R CC and L/R MLO) are processed by a shared spatio-temporal feature extraction module (STFE) consisting of a ResNet-18 backbone and a temporal encoder to model longitudinal dependencies. For the primary task, (A) cancer classification integrates (i) a suspicious-area extraction branch and a Mask Feature Learner (MFL) for lesion-focused refinement and (ii) a Multi-Scale Feature Fusion module (MSFF) to capture short-, mid-, and long-term temporal patterns. The auxiliary modules leverage intermediate features to compute breast symmetry scores (B) and estimate tissue density (C). All three tasks are optimized jointly through an adaptive weighting strategy. In addition, an Independent Laterality Classifier (D) is introduced to predict tumor laterality for cancer-positive cases. This module is trained separately using only the cancer-positive samples and leverages spatial features extracted from the last layer of the STFE.

#### 1) Cancer classification task

As shown in **Fig. 1(A)**, The cancer classification branch consists of three major components: the Spatial-Temporal Feature Extraction (STFE) module, the Multi-Scale Feature Fusion (MSFF) module, and the Mask Feature Learner (MFL).

##### Spatial-Temporal Feature Extraction module

The STFE module extracts view-specific spatial features and models longitudinal dependencies across serial mammograms. Each bilateral composite image is processed by pretrained ResNet-18 backbone [24] to obtain spatial feature representations. These features are then flattened into sequences and passed through Transformer encoders to capture temporal changes across examinations:

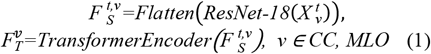

The resulting view-specific temporal representations, 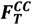 and 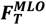, encode longitudinal changes in breast tissue and are used as inputs to the subsequent MSFF module. In addition to the final spatial features used for temporal modeling, intermediate feature maps extracted from the second layer of ResNet-18, denoted as 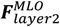 and 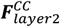 , are provided to the symmetry-analysis and breast-density-classification branches. The detailed processing of these intermediate features is described in the following sections.

##### Multi-Scale Feature Fusion module

The MSFF module integrates temporal information at multiple scales and fuses complementary information from CC and MLO views. To capture temporal patterns over different effective intervals, three parallel one-dimensional temporal convolutions with kernel sizes of 1, 3, and 5 are applied to the view-specific temporal features. These operations generate short-, intermediate-, and long-range temporal representations for each view. For each temporal scale, a cross-view attention mechanism is used to fuse the corresponding CC and MLO features. This allows the model to adaptively weight complementary information from the two mammographic projections. The scale-specific fused representations are then aggregated to generate the final global longitudinal representation:

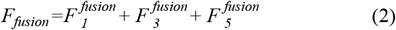

This design enables the cancer classification branch to jointly capture longitudinal tissue evolution, multi-view information, and temporal-scale-dependent changes.

##### Mask Feature Learner module

The MFL module incorporates localized suspicious-region information into the cancer classification branch. A pretrained weakly supervised mammography model is first used to generate saliency maps highlighting image regions with a higher likelihood of abnormal findings [25]. These saliency maps are then processed by a lightweight convolutional network followed by global average pooling to generate a compact suspicious-region representation, ***F***_***mask***_. The suspicious-region representation is integrated with the global longitudinal feature representation using element-wise multiplication:

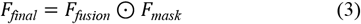

where ⊙ denotes element-wise multiplication. This operation acts as an implicit attention mechanism that emphasizes image regions more relevant to cancer prediction while preserving global longitudinal context. The resulting representation, *FF*_*final*_, is passed through a dropout layer and a fully connected classification head to generate the final cancer-classification logits.

#### 2) Symmetry analysis task

As shown in **Fig. 1(B)**, the symmetry analysis branch was designed to encourage the model to learn bilateral asymmetry features that may be relevant to malignancy. Intermediate spatial features from the second ResNet-18 layer 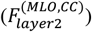 are refined using a Convolutional Block Attention Module (CBAM) [26], which applies channel and spatial attention to emphasize informative breast tissue regions. The refined feature maps are horizontally split into left and right breast components, followed by global average pooling to generate left- and right-breast feature vectors. Bilateral symmetry is quantified using cosine similarity between the left and right feature vectors for both CC and MLO views. The symmetry score is averaged across time points, and a symmetry loss is applied to cancer-positive cases to encourage sensitivity to pathological asymmetry:

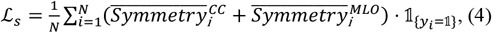

where ***N*** is the batch size, *YY*_*ii*_ is the cancer label, and 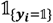 indicates cancer-positive cases.

#### 3) Breast density classification task

As shown in **Fig. 1(C)**, the breast density classification branch uses intermediate features 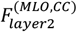 shared with the main cancer classification network. We propose a multi-scale Breast Density Feature Extraction module (BDFE) that integrates Adaptive Spatial Pyramid Pooling (ASPP) and self-attention mechanisms to achieve adaptive weighting and fusion of breast density features at different scales. This module effectively captures global and multi-scale breast parenchymal patterns. Specifically, the module uses a 1×1 convolution for feature transformation, followed by adaptive spatial pyramid pooling with multiple pooling scales to summarize breast tissue distribution at different spatial resolutions. The resulting multi-scale density features are concatenated and refined using fully connected layers and channel reweighting. A task-specific self-attention module is then used to model temporal dependencies in breast density features across longitudinal examinations. This branch was designed separately from the cancer classification temporal module because breast density changes generally follow a slower and more global temporal pattern than lesion-related morphological changes.

#### 4) Laterality classification task

As shown in **Fig. 1(D)**, the independent laterality classifier (ILC) provides an interpretable localization output for cancer-positive cases with available left/right tumor laterality labels. This branch uses spatial features extracted from the final convolutional block of the main cancer classification network. After global average pooling, the feature vector is passed through a dropout layer and a fully connected layer to predict tumor laterality. The laterality classifier is trained independently using cancer-positive cases only and is not included in the main multi-task loss. During inference, the laterality output is interpreted only for cases classified as cancer-positive by the primary cancer classification branch. This design preserves task specificity while providing an additional anatomical localization output.

### D. Loss Function Design

The cancer-classification, breast-density, and symmetry-analysis tasks were jointly optimized using a dynamically weighted multi-task loss. Learnable log-scale parameters ***W***_log_ = [log *w*_*c*_ , log *w*_*d*_ , log *w*_*s*_] , were transformed into normalized task weights using a temperature-scaled SoftMax:

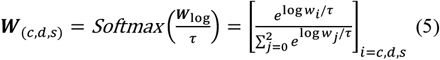

where τ is a parameter controlling the smoothness of the weight distribution. The joint loss was defined as

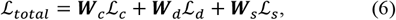

where ℒ_*c*_ and ℒ_*d*_ denote the cancer- and density-classification losses, respectively, and ℒ_*s*_ is the symmetry loss defined in (4). Both classification tasks were optimized using cross-entropy loss. To stabilize the learned task weights, a regularization term was added:

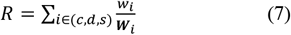

The final dynamic loss was therefore:

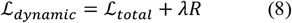

where λ controls the contribution of the regularization term. This strategy allows the relative importance of each task to be adjusted during training rather than fixed manually.

The laterality classifier was trained independently because laterality labels were available only for cancer-positive cases. It was therefore excluded from the dynamic multi-task objective and optimized separately using a binary classification loss.

## III. EXPERIMENTS DESIGN

### A. Comparative Models

We compared MuSTAF with two groups of models. First, architecture-level baselines used the same preprocessed longitudinal multi-view inputs and cross-validation partitions. These baselines combined ResNet-18 with Transformer, long short-term memory, recurrent neural network, temporal convolutional network, or three-dimensional convolutional neural network modules [27-31]. Each baseline used task-specific heads for cancer and density classification.

Second, we compared MuSTAF with three published MG AI models: Mirai [32], LRP-NET [15] and GMIC [33] These models were adapted to the common endpoint of individual-level binary cancer discrimination. Mirai was evaluated using its 1-year risk score as a continuous prediction score. LRP-NET used up to the three most recent screening examinations per individual. GMIC generated image-level cancer scores that were aggregated to the individual level. Internal validation used held-out predictions from the 7-fold cross-validation setting when applicable. External validation was performed on CSAW-CC without using external images for training, hyperparameter tuning, threshold selection, or checkpoint selection. Because benchmark models did not provide the same auxiliary outputs as MuSTAF, comparisons focused on cancer classification.

### B. Ablation Experiments and Model Explainability

Ablation experiments used the same individual-level cross-validation partitions as the primary MuSTAF model. We first evaluated task-level ablations by removing breast density supervision or symmetry-informed learning. We then compared longitudinal input lengths using the three most recent examinations versus the five most recent examinations. Finally, we performed module-level ablations by removing the MSFF module, MFL module and auxiliary task modules.

Model explainability was assessed qualitatively using class activation maps from the cancer classification branch. Maps were overlaid on representative mammograms to assess whether model attention corresponded to clinically relevant regions. Laterality predictions were evaluated in cancer-positive cases with available left/right tumor labels.

### C. Evaluation Metrics and Statistical Analysis

Model discrimination was assessed using AUC, Accuracy (ACC), F1 score, and confusion matrices were calculated at a fixed probability threshold of 0.5 unless otherwise specified. Internal validation metrics were computed for each held-out fold and summarized as mean ± standard deviation. External AUC and accuracy 95% confidence intervals were estimated using nonparametric individual-level bootstrap resampling. Comparisons with baseline, ablation, and benchmark models used individual-level predictions. Explainability was assessed qualitatively using representative visualization examples.

## IV. Results

### A. Internal Validation and Baseline Comparison

In individual-level 7-fold cross-validation, MuSTAF achieved the best overall performance among the evaluated architecture-level baseline models (**Table 2A**). For cancer classification, MuSTAF achieved an AUC of 0.84 ± 0.07, compared with 0.76 ± 0.07 for the best baseline model, ResNet-18+Transformer. Across the seven held-out folds, MuSTAF correctly classified 136 of 176 cancer cases and 147 of 175 non-cancer controls. This corresponded to a sensitivity of 77.3%, specificity of 84.0%, accuracy of 80.6%, and F1 score of 80.0%. For breast density classification, MuSTAF achieved an AUC of 0.83 ± 0.08. The model correctly classified 158 of 185 dense cases and 119 of 166 non-dense cases, with sensitivity of 85.4%, specificity of 71.7%, accuracy of 79.7%, and F1 score of 81.9%. This exceeded the best architecture-level baseline for density classification, ResNet-18+TCN, which achieved an AUC of 0.79 ± 0.08. Models with explicit temporal modeling generally outperformed the 3D-CNN baseline.

**TABLE II.** Performance Comparison of Baseline Models and Mustaf Ablation Variants for Cancer and Breast Density Classification.

| Model | Cancer classification |  |  | Density classification |  |  | Param (M) |
| --- | --- | --- | --- | --- | --- | --- | --- |
|  | ACC mean (STD) | AUC mean (STD) | F1 Score mean (STD) | ACC mean (STD) | AUC mean (STD) | F1 Score mean (STD) |  |
| A. Architecture-level baselines |  |  |  |  |  |  |  |
| ResNet-18+Transformer | 0.756 (0.065) | 0.754 (0.064) | 0.748 (0.060) | 0.781 (0.049) | 0.776 (0.047) | 0.779 (0.055) | 22.3 |
| ResNet-18+LSTM | 0.731 (0.088) | 0.763 (0.090) | 0.752 (0.091) | 0.762 (0.079) | 0.731 (0.072) | 0.741 (0.098) | 11.5 |
| ResNet-18+TCN | 0.736 (0.084) | 0.738 (0.086) | 0.732 (0.085) | 0.786 (0.108) | 0.791 (0.075) | 0.781 (0.120) | 10.5 |
| ResNet-18+RNN | 0.744 (0.041) | 0.746 (0.043) | 0.741 (0.042) | 0.761 (0.096) | 0.754 (0.070) | 0.748 (0.111) | 11.8 |
| 3D-CNN | 0.681 (0.045) | 0.676 (0.046) | 0.683 (0.048) | 0.744 (0.041) | 0.736 (0.036) | 0.738 (0.053) | 46.5 |
| B. Task-level ablation |  |  |  |  |  |  |  |
| w/o Density classification task | 0.773 (0.036) | 0.773 (0.035) | 0.771 (0.037) | - | - | - | 26.0 |
| w/o Symmetry Analysis | 0.776 (0.097) | 0.780 (0.103) | 0.773 (0.092) | 0.7647 (0.0948) | 0.754 (0.092) | 0.763 (0.087) | 29.1 |
| C. Module-level ablation |  |  |  |  |  |  |  |
| w/o Transformer Encoder | 0.766 (0.033) | 0.765 (0.032) | 0.764 (0.034) | 0.818 (0.087) | 0.815 (0.067) | 0.811 (0.096) | 17.2 |
| w/o MSFF | 0.775 (0.027) | 0.781 (0.034) | 0.761 (0.011) | 0.696 (0.090) | 0.744 (0.065) | 0.715 (0.078) | 27.4 |
| w/o MFL | 0.757 (0.024) | 0.756 (0.027) | 0.754 (0.027) | 0.707 (0.111) | 0.749 (0.112) | 0.670 (0.146) | 28.2 |
| w/o CBAM | 0.769 (0.050) | 0.781 (0.052) | 0.752 (0.065) | 0.781 (0.078) | 0.786 (0.037) | 0.744 (0.104) | 29.4 |
| w/o BDFE | 0.796 (0.024) | 0.797 (0.039) | 0.794 (0.031) | 0.773 (0.043) | 0.780 (0.047) | 0.784 (0.035) | 23.1 |
| Full MuSTAF (ours) | 0.806 (0.065) | 0.843 (0.043) | 0.801 (0.074) | 0.797 (0.104) | 0.833 (0.082) | 0.819 (0.041) | 29.7 |

### B. Ablation studies of Multi-task Learning, Longitudinal modeling and Model Components

We conducted three complementary sets of ablation experiments. In the task-level ablations (**Table 2B**), an entire auxiliary task and its corresponding supervision were removed from joint optimization. The longitudinal analyses evaluated the contribution of temporal modeling by removing the Transformer encoder and by comparing different longitudinal input windows. In contrast, the module-level ablations (**Table 2C**) retained the associated prediction task and loss function but removed a specific architectural component used to learn its representation.

Task-level ablations showed that auxiliary learning contributed to cancer classification (**Table 2B**). Removing breast density classification reduced cancer AUC from 0.84 to 0.77. Removing symmetry-informed learning reduced cancer AUC to 0.78. Symmetry removal also reduced density classification AUC from 0.83 to 0.75, suggesting complementary information from density- and symmetry-related representations.

Module-level ablations showed that key MuSTAF components contributed non-redundant information (**Table 2C**). Removing the MSFF module produced the largest decrease in density classification AUC, from 0.83 to 0.74. Removing the mask feature learner reduced cancer classification AUC to 0.76. For modules related to symmetry analysis, removing the CBAM, which models bilateral breast symmetry, led to a notable decline in cancer classification performance (AUC from 0.84 to 0.78), underscoring the significance of symmetry cues in distinguishing malignant from benign cases. Lastly, eliminating the BDFE slightly reduced the accuracy and F1-score of density classification, suggesting its utility in modeling global tissue distribution patterns.

The contribution of longitudinal information was evaluated from both architectural and input-window perspectives. Removing the Transformer encoder reduced the cancer-classification AUC from 0.84 to 0.77, indicating that explicit modeling of temporal dependencies across serial examinations contributed to cancer discrimination. We further compared models using the three versus five most recent mammographic examinations. The three-examination setting achieved a higher internal cancer-classification AUC than the five-examination setting (0.84 vs 0.80) and also showed better external performance (0.72 vs 0.66). These findings supported the use of the three most recent examinations as the primary longitudinal input window.

### C. Benchmark Comparison and External Validation

We compared MuSTAF with three published MG AI models, including LRP-NET [15], GMIC [33], and Mirai [32], using individual-level cancer classification as the common evaluation task. LRP-NET incorporates longitudinal mammographic information and outputs an individual-level Wnear-term cancer risk score. GMIC is a single-exam lesion-aware MG model that outputs cancer prediction scores with lesion-attention information. Mirai is a single-exam MG AI model that outputs multi-year risk estimates. Its 1-year risk score was used as an adapted individual-level cancer classification score. Because these benchmark models do not provide multi-task outputs, the comparison focused on cancer classification.

The result shown in **Fig. 2**, in the in-house cohort, MuSTAF achieved the strongest performance among the published benchmark models, with an AUC of 0.84 (95% CI: 0.78–0.90) and accuracy of 0.81 (95% CI: 0.76–0.84). LRP-NET achieved an AUC of 0.74 (95% CI: 0.66–0.81) and accuracy of 0.71 (95% CI: 0.65–0.77). GMIC achieved an AUC of 0.71 (95% CI: 0.63–0.79) and accuracy of 0.64 (95% CI: 0.54–0.73). Mirai achieved an AUC of 0.81 (95% CI: 0.78–0.83) and accuracy of 0.73 (95% CI: 0.69–0.75). These results suggest that MuSTAF and Mirai had the strongest in-house performance, while MuSTAF outperformed the longitudinal LRP-NET model and the single-exam GMIC model.

**Fig 2.**
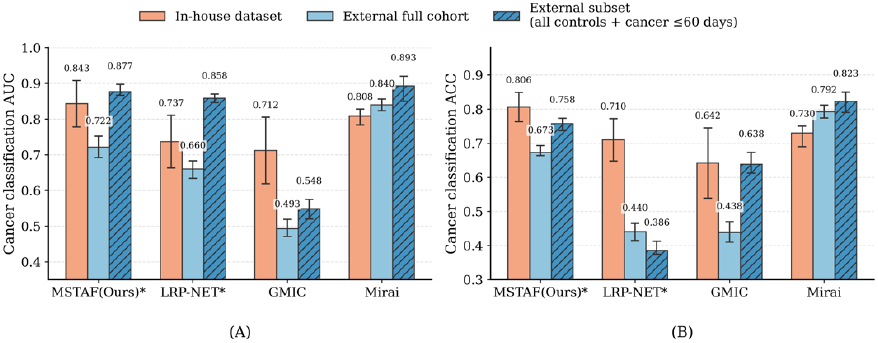
Benchmark comparison and external validation. (A) Cancer classification AUC and (B) ACC of MuSTAF, LRP-NET, GMIC, and Mirai on the in-house cohort (orange), the full external CSAW-CC cohort (light blue), and the timing-restricted subset of CSAW-CC cohort (dark blue with shadow). Error bars indicate standard deviation for internal cross-validation and 95% confidence intervals for external validation.

On the full CSAW-CC external validation cohort, MuSTAF outperformed LRP-NET and GMIC, although its performance decreased relative to the in-house cohort. The external AUCs were 0.72 (95% CI: 0.69–0.75) for MuSTAF, 0.66 (95% CI: 0.63–0.68) for LRP-NET, and 0.49 (95% CI: 0.47–0.51) for GMIC. Mirai maintained strong external performance, with an AUC of 0.84 (95% CI: 0.82–0.85), broadly consistent with its previously reported 1-year risk prediction performance on CSAW-CC cohort from Karolinska University Hospital [32].

We further investigated the potential causes of performance reduction of MSTAF, LRP-NET and GMIC on the external cohort (see more details in sections below). Most importantly, the cancer cases in the external cohort have substantially longer time intervals between the latest screening examination and the cancer diagnosis (60% < 60 days, 24.9% 60-729 days, 15.1% > 730 days) than our in-house cohort (94.6% < 60 days, 5.4% 60-729 days). Therefore, we evaluated a temporally aligned external subset that included all controls and only cancer cases diagnosed within 60 days of the corresponding mammographic examination, so to reduce the temporal mismatch between the mammographic evidence and the cancer-classification endpoint. In this subset, the AUC increased from 0.722 to 0.877 for MuSTAF and from 0.660 to 0.858 for LRP-NET. More modest increases were observed for GMIC, from 0.493 to 0.548, and for Mirai, from 0.840 to 0.893. MuSTAF accuracy also increased from 0.673 to 0.758, while GMIC and Mirai accuracies increased from 0.438 to 0.638 and from 0.792 to 0.823, respectively. LRP-NET achieved an accuracy of 0.386 in the temporally aligned subset despite its improved AUC. Overall, MuSTAF and Mirai showed the strongest discrimination in the temporally aligned external analysis, and the marked AUC improvements for MuSTAF and LRP-NET indicate that the interval between imaging and cancer diagnosis was an important contributor to external performance.

### D. Laterality Classification and Model Interpretability

The independent laterality classifier was trained only on in-house cancer-positive cases (n = 176). It achieved an AUC of 0.80, accuracy of 78%, and F1 score of 0.78. To further interpret the model’s spatial reasoning, we examined the alignment between attention regions and ground-truth tumor laterality. In particular, we conducted a Grad-CAM–based visualization analysis with the final convolutional layer of the ResNet-18 backbone in the ILC module, showing attention patterns overlapping with clinically relevant breast regions. As shown in **Fig. 3**, we present representative Grad-CAM heatmaps overlaid on longitudinal mammographic views. Warmer regions indicate stronger model attention. In examples such as **Fig. 3A**, the model accurately localizes the malignant lesion on the correct side. In benign cases (**Fig. 3B**), attention is more spatially diffuse, yet still reflects plausible anatomical priors consistent with radiological patterns.

**Fig 3.**
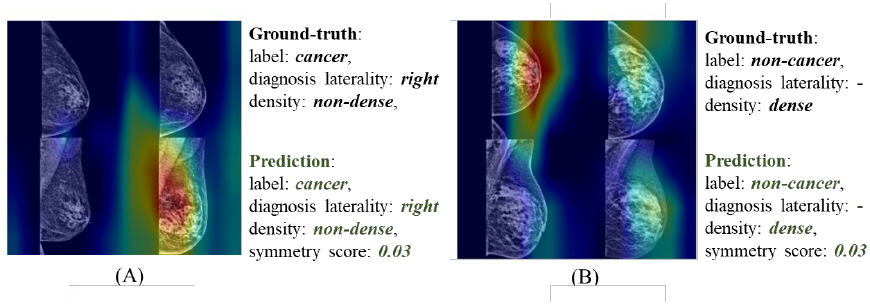
Grad-CAM heatmaps and model outputs from MuSTAF. Grad-CAM visualizations derived from the final convolutional layer of the ResNet-18 backbone, overlaid on longitudinal mammographic images. Warmer regions indicate stronger model attention. Each panel displays the model’s predictions alongside the ground-truth diagnosis, including cancer status, tumor laterality, breast density, and symmetry score.

### E. Dataset Shift Analysis

To systematically investigate factors underlying the external generalization gap, we performed a dataset-shift analysis comparing the in-house and external cohorts. The motivation for this analysis was that cancer classification performance decreased in the external cohort, suggesting that the model retained transferable representations of global breast parenchymal patterns but remained vulnerable to shifts affecting cancer-specific decision signals. We therefore examined potential differences between cohorts at the level of longitudinal examination structure, and model sensitivity to controlled image-level perturbations. This analysis was to determine whether the observed external performance gap was more consistent with reduced longitudinal evidence, appearance/texture shift, or simple geometric inconsistency.

#### 1) Longitudinal Structure Differences Between Cohorts

We compared the longitudinal structure of the in-house and external cohorts because MuSTAF explicitly models temporal information across prior mammographic examinations. As shown in **Fig. 4**, the two cohorts differed substantially in the availability of longitudinal examinations. In the overall cohort, the external dataset contained a higher proportion of cases with only one or two available examinations, whereas the in-house cohort had a larger fraction of cases with four or five examinations (**Fig. 4A**). This indicates that, although the external cohort was larger in sample size, many external cases provided a shorter effective imaging history for temporal modeling. We also compared follow-up span, defined as the time difference between the earliest and latest available examinations for each patient (**Fig. 4B**). Cancer-positive external cases showed a larger fraction of zero-year or short follow-up spans, consistent with the higher prevalence of single-exam or short-sequence cases. By comparison, the in-house cancer cohort more frequently showed multi-year longitudinal coverage. Thus, the temporal model has less longitudinal evidence available for many external patients.

**Fig 4.**
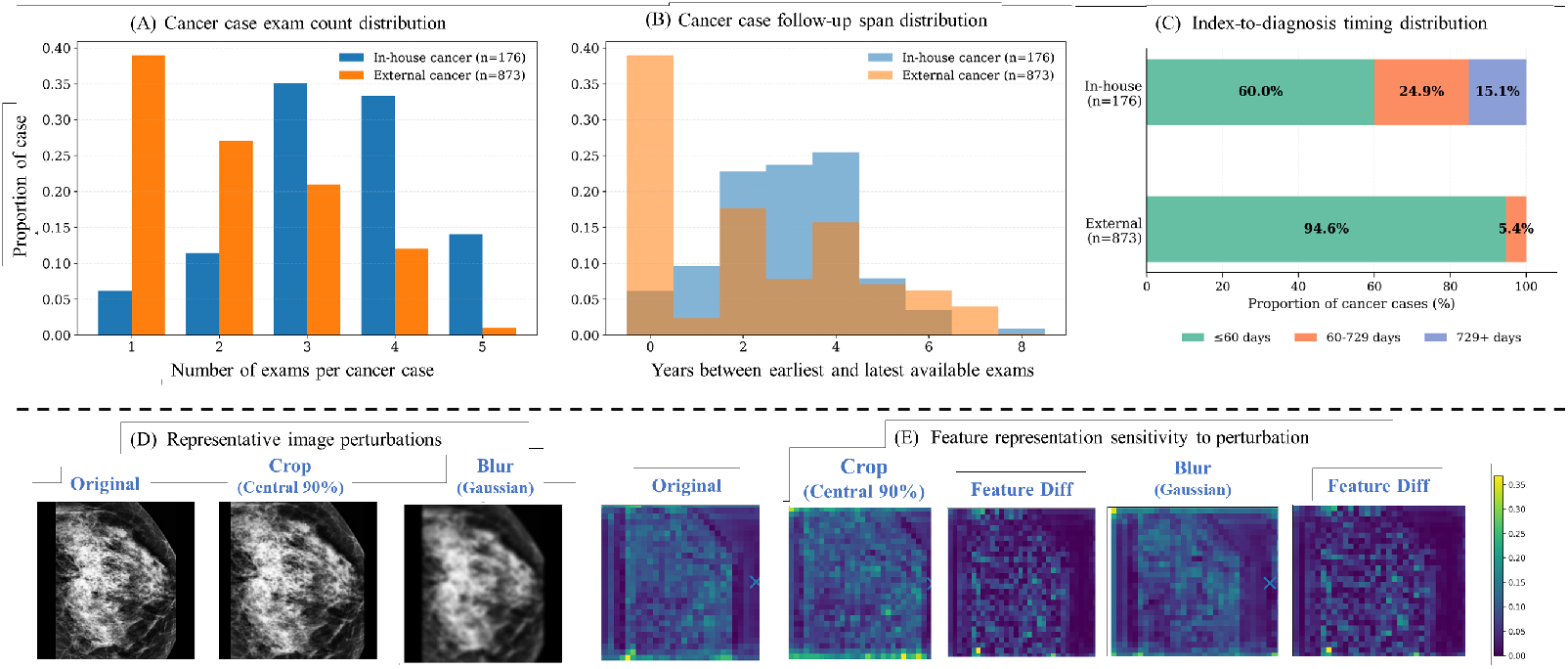
(A–C) Comparison of longitudinal data structure between the in-house and external cohorts among cancer-positive cases: (A) number of available mammographic examinations, (B) longitudinal follow-up span, and (C) timing of the index examination relative to cancer diagnosis. The external cohort contained a larger proportion of short-sequence cases, indicating less longitudinal progression and stability information for temporal modeling. (D) Representative perturbations applied to a single mammographic view while all other views and time points were held unchanged, including central cropping at 90% of the original field of view and Gaussian blurring. (E) Representative backbone activation maps before and after perturbation, illustrating perturbation-induced changes in learned feature representations

In addition, we examined the temporal proximity of the selected latest cancer-positive examination to diagnosis (**Fig. 4C**). This analysis reflected the timing structure available within each cohort. In the external CSAW-CC cohort, 60.0% of selected index cancer-positive examinations were obtained within 60 days of diagnosis, whereas 24.9% were obtained 60-729 days before diagnosis and 15.1% were obtained 730 days or more before diagnosis. In contrast, the in-house cohort was predominantly composed of near-diagnosis examinations, with 94.6% obtained within 60 days and 5.4% obtained 60-729 days before diagnosis. Thus, even the latest screening examination was always used as the index examination, the external cohort contained a broader mixture of near-diagnosis and earlier prior examinations. This sequence-level mismatch is observed to be one of the biggest contributors to the observed external cancer performance gap as reported above. In fact, when the latest examination is years before the cancer diagnosis, the cancer classification task in fact becomes a multi-year risk prediction task, which is matching better with the design of risk assessment model (such as Mirai), which also explains why Miria performance did not show an obvious drop as other model in the whole CSAW-CC cohort. These observations indicate that for AI models trained for cancer detection on the dataset with a strong correspondence between the visible mammographic findings and immediate cancer incidence, it is important to keep the generalized model applications within the immediate cancer detection to maintain a reliable performance.

#### 2) Input Resolution and Preprocessing Sensitivity

We further examined the effect of input resolution and preprocessing on comparator model performance. In the initial harmonized benchmark setting, all models were evaluated using the same resized 224 × 224 mammographic inputs to match the MuSTAF input size. This strategy provided a common input format across models, but it did not match the native input setting of Mirai [12], which was designed for higher-resolution mammographic images and model-specific preprocessing. Under the 224 × 224 downsampled input setting, Mirai showed substantially reduced external performance on CSAW-CC, with an AUC of 0.50 (95% CI: 0.47–0.51). After using a preprocessing pipeline more consistent with Mirai’s native input requirements and preserving original presentation mammographic resolution, Mirai achieved the external AUC of 0.840 (95% CI: 0.823–0.856) reported above. This marked difference indicates that pretrained single-time-point mammography models can be highly sensitive to input resolution and preprocessing. Because these models rely primarily on the current examination rather than longitudinal change, preservation of fine mammographic details may be particularly important for capturing subtle cancer-related findings, including calcifications, architectural distortion, and focal asymmetry. Therefore, model-specific preprocessing requirements should be considered when interpreting cross-model benchmark comparisons and external validation results.

#### 3) Perturbation Assessment of Image-level Domain Shift

To further evaluate whether image-level differences could contribute to the external performance gap, we conducted a controlled perturbation analysis to characterize the model’s sensitivity to specific types of image variation. Rather than directly altering the full longitudinal sequence, we applied perturbations to a single target mammographic view from one examination while keeping all other views and time points unchanged. This design allowed us to isolate the effect of each image-level perturbation on both the model output and intermediate feature representations. As shown in **Fig. 4D**, the tested perturbations included cropping and Gaussian blurring. The perturbation analysis showed that the cancer prediction branch was particularly sensitive to changes affecting image texture and effective resolution. Among the tested perturbations, downsampling, strong blurring, and marked tightening of the field of view produced the largest changes in predicted cancer probability and intermediate feature representations.

We also examined perturbation-induced changes in feature representations. For representative cases, activation maps from the MuSTAF convolutional backbone were compared before and after perturbation. As shown in **Fig. 4E**, blur and crop altered spatial activation patterns, with visible changes in the activation-difference maps. These visual changes were consistent with layer-wise quantitative feature-drift analysis, which showed that perturbation effects were more pronounced in the temporal encoder than in early convolutional features. In this representative case, blurring and cropping produced cosine distances of 0.173 and 0.101, respectively, in the final Transformer encoder layer, whereas the corresponding backbone-layer distances were less than 0.001. Final cancer-probability changes were smaller, with a maximum absolute change of 0.0027, suggesting that image-level perturbations can substantially alter internal temporal representations even when output probabilities remain relatively stable.

## V. Discussion

In this study, we developed and evaluated MuSTAF, a longitudinal multi-view multi-task deep learning framework for patient-level breast cancer classification from screening MG. MuSTAF integrates serial bilateral CC and MLO examinations, temporal fusion, mask-guided suspicious-region refinement, and auxiliary learning from breast density and bilateral symmetry. In internal validation, MuSTAF outperformed architecture-level baselines and showed strong performance compared with published MG AI models. In external validation on CSAW-CC, MuSTAF remained the best-performing longitudinal model, although Mirai achieved the highest overall external performance. These findings suggest that longitudinal multi-view modeling and clinically aligned multi-task supervision can improve cancer discrimination. They also show that external performance depends on the temporal structure and task definition of the validation cohort.

A key strength of MuSTAF is that its auxiliary tasks reflect routine mammographic interpretation. Radiologists assess not only suspicious focal findings, but also breast density, bilateral symmetry, and lesion laterality. Breast density is related to masking and cancer risk. Bilateral comparison helps identify unilateral or developing abnormalities. Tumor laterality provides an anatomical localization output for cancer-positive predictions. By jointly learning these outputs, MuSTAF captures both focal lesion-related features and broader contextual information. The ablation experiments supported this design, as removing density or symmetry supervision reduced primary cancer classification performance. These auxiliary outputs also provide testable checkpoints for human-in-the-loop evaluation and help improve radiologist trust.

The longitudinal input analysis showed that using the three most recent screening examinations performed better than five examinations. This suggests that very old prior mammograms may introduce noise rather than useful diagnostic information. Breast tissue changes are not necessarily linear over time, and remote examinations may be less relevant to the current cancer outcome. This finding has practical implications, as shorter longitudinal windows reduce model complexity, lower computational and storage demands, and make external validation easier in cohorts with incomplete imaging histories. Future longitudinal MG models should therefore optimize not only how temporal information is modeled, but also how much prior imaging history is used.

The benchmark comparison highlights differences among current MG AI strategies. GMIC [33] is a single-exam lesion-aware classification model. LRP-NET [15] incorporates longitudinal mammographic information for near-term risk prediction. Mirai is considered a state-of-the-art (SOTA) deep learning model for breast cancer risk prediction and is being evaluated in ongoing trials (e.g. MyPeBS [34]) to guide screening recommendations. In the in-house cohort, MuSTAF achieved the strongest performance among the evaluated models, which is consistent with its design. Compared with GMIC and Mirai, MuSTAF explicitly models serial examinations. Compared with LRP-NET, MuSTAF further incorporates density supervision, bilateral symmetry learning, and CC/MLO cross-view fusion.

The external CSAW-CC results provide a more nuanced interpretation. MuSTAF outperformed LRP-NET and GMIC, but all three dropped clearly, and Mirai became the top performer. The main driver of this drop appears to be the interval between the latest screening and diagnosis. In CSAW-CC, many cancer-positive cases had a multi-year interval between their latest screening and cancer diagnosis, where the mammogram may contain weaker or less visible cancer-related findings. For these cases, the evaluated task therefore becomes closer to risk prediction than immediate cancer classification. This setting is better aligned with Mirai’s original design.

The timing-restricted subset analysis supports this interpretation. When cancer-positive cases were limited to those screened within 60 days before diagnosis, while all non-cancer controls were retained, MuSTAF, LRP-NET, and GMIC showed significant improvment. This suggests that reducing temporal mismatch between the cancer label and visible mammographic evidence brought the evaluation back to the detection task these models were trained for, thus recovering the performance accordingly.

Timing was not the only factor affecting generalization. The domain-shift analysis also showed effects related to longitudinal structure mismatch, scale and field-of-view mismatch, image-level appearance differences, and texture domain shift. In addition, benchmark performance was sensitive to input resolution. This suggests that spatial downsampling to 224 × 224 in longitudinal models preprocessing may remove fine mammographic details relevant to cancer discrimination, including subtle calcifications, architectural distortion, and focal asymmetry.

Overall, we identified several opportunities to further strengthen and extend this work. First, CSAW-CC provided independent external validation, but additional multi-institutional testing is needed across vendors, acquisition protocols, and screening populations. Public longitudinal MG datasets remain limited, and consortium-level studies will be important for validating temporal screening AI models. Second, benchmark comparison was limited by differences in model objectives, input requirements, and implementation availability. For example, Mirai was adapted from risk prediction by using its 1-year risk score, whereas MuSTAF was trained directly for cancer classification with auxiliary tasks. Third, MuSTAF used saliency maps generated by a pretrained MG detection model, which may introduce dependence on that model. Future work could incorporate lesion localization into an end-to-end framework. A high-resolution or multi-resolution image branch may also help preserve subtle local findings while retaining longitudinal temporal modeling. Finally, although the source screening cohort was prospectively collected, this analysis was retrospective and used a nested case-control design. This does not reflect cancer prevalence in population screening. Prospective reader studies in full screening cohorts are warranted to evaluate the translational benefits of MuSTAF regarding improving recall rate, positive predictive value, sensitivity, specificity, reading time, radiologist performance, clinical decision-making, and overall workflow impact.

## VI. CONCLUSION

MuSTAF demonstrated the potential of longitudinal multi-view multi-task learning for breast cancer detection from screening mammography. The framework integrated temporal imaging patterns and produced auxiliary outputs aligned with mammographic interpretation, including breast density, bilateral symmetry, and tumor laterality. A three-examination input window provided the most favorable performance. Prospective multicenter validation and reader studies are needed to determine clinical utility in routine screening.

## Data Availability

All data produced in the present study are available upon reasonable request to the authors

